# Interpretable Clinical Genomics with a Likelihood Ratio Paradigm

**DOI:** 10.1101/2020.01.25.19014803

**Authors:** Peter N. Robinson, Vida Ravanmehr, Julius O.B. Jacobsen, Daniel Danis, Xingmin Aaron Zhang, Leigh C. Carmody, Michael Gargano, Courtney L. Thaxton, UNC Biocuration Core, Justin Reese, Manuel Holtgrewe, Sebastian Köhler, Julie A. McMurry, Melissa A. Haendel, Damian Smedley

## Abstract

Human Phenotype Ontology (HPO)-based analysis has become standard for genomic diagnostics of rare diseases. Current algorithms use a variety of semantic and statistical approaches to prioritize the typically long lists of genes with candidate pathogenic variants. These algorithms do not provide robust estimates of the strength of the predictions beyond the placement in a ranked list, nor do they provide measures of how much any individual phenotypic observation has contributed to the prioritization result. However, given that the overall success rate of genomic diagnostics is only around 25–50% or less in many cohorts, a good ranking cannot be taken to imply that the gene or disease at rank one is necessarily a good candidate. Likelihood ratios (LR) are statistics for summarizing diagnostic accuracy, providing a measure of how much more (or less) a patient with a disease has a particular test result compared to patients without the disease. Here, we present an approach to genomic diagnostics that exploits the LR framework to provide an estimate of (1) the posttest probability of candidate diagnoses; (2) the LR for each observed HPO phenotype, and (3) the predicted pathogenicity of observed genotypes. LIkelihood Ratio Interpretation of Clinical AbnormaLities (LIRICAL) placed the correct diagnosis within the first three ranks in 92.9% of 384 cases reports comprising 262 Mendelian diseases, with the correct diagnosis having a mean posttest probability of 67.3%. Simulations show that LIRICAL is robust to many typically encountered forms of genomic and phenomic noise. In summary, LIRICAL provides accurate, clinically interpretable results for phenotype-driven genomic diagnostics.

---

Phenotype-driven prioritization of candidate genes and diseases is a well-established approach towards genomic diagnostics in rare disease. ^1–12^ Most current approaches use the Human Phenotype Ontology (HPO) for annotating the set of phenotypic abnormalities observed in the individual being investigated by exome or genome sequencing (WES/WGS). The HPO contains 14,813 terms arranged as a directed acyclic graph in which edges represent subclass relations; 13,182 of these terms represent phenotypic abnormalities. For instance, `Abnormal renal cortex morphology` (HP:0011035) is a subclass of `Abnormal renal morphology` (HP:0012210). The HPO project additionally provides computational disease models of 7623 rare diseases that are constructed from HPO terms and metadata that define the diseases based on the phenotypic abnormalities that characterize them, their modes of inheritance, and in many cases the age of onset of diseases or phenotypic features and the overall frequencies of features in a disease. ^13^ For instance, Meckel syndrome type 7 is characterized by `Patent ductus arteriosus` (HP:0001643) with a frequency of two of seven patients and `Antenatal onset` (HP:0030674). ^14^

WES/WGS typically reveals tens or hundreds of variants that are predicted to be deleterious by common computational frameworks, and therefore the analysis of such data generally requires some additional criterion to prioritize genes. ^15^ Phenotypic approaches leverage the proband’s observed phenotypic abnormalities to assess candidate diseases by searching diseases with similar phenotypic abnormalities that are associated with genes that harbor a predicted pathogenic variant. ^16^ However, current algorithms for phenotype-driven genomic diagnostics have a number of shortcomings that represent impediments to the successful implementation of genomic testing outside of specialist centers. All current approaches that we are aware of present their results as an ordered list of candidate genes or diseases. The overall sucess rate of genomic diagnostics depends on the cohort and the NGS technique, but the overall rate is still hovering at about 40% for a wide range of conditions. ^17–20^ Therefore, one must expect that, in many cases, the top-ranked gene is actually not a good candidate. Also, existing approaches do not provide a framework for deciding how many candidates in the ranked list are worthy of detailed examination. Therefore, it would be desirable to provide a transparent measure of how good the top predictions are, and why. Such an approach could reduce the number of candidates that busy diagnostic labs have to review. Finally, current approaches do not provide information about how much individual phenotypic features contribute to the computational prediction. For clinical use, approaches that allow users to understand the reasons for the computational predictions are preferable to black-box algorithms and better support clinical decision making. ^21^

## Results

In this work, we present an approach that addresses the aforementioned shortcomings by applying the likelihood ratio (LR) framework to phenotype-driven genomic diagnostics. The LR is defined as the probability of a given test result in an individual with the target disorder divided by the probability of that same result in an individual without the target disorder. The LR framework allows multiple test results to be combined by multiplying the individual ratios, and also relates the pretest probability to the posttest probability in a way that can be used to guide clinical decision making. ^22–24^

### The LIRICAL algorithm

We define a LR-based model of the clinical examination of a patient being investigated for a suspected but unknown Mendelian disorder as follows. Each recorded phenotypic observation is defined as a clinical test. The probability of a person with disease 𝒟 having a phenotypic abnormality encoded by HPO term *h*_*i*_, denoted as 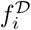, is taken to be the frequency with which the abnormality is observed in affected individuals as recorded in the computational disease models of the HPO project based on literature biocuration (a default value of 100% is used if specific frequency information is not available). For many diseases and features, an overall frequency of the feature is known; for instance, 19/437 persons (*∼* 4%) with neurofibromatosis type 1 have seizures. ^25^ On the other hand, 338/442 individuals (*∼*87%) with this disease have multiple café-au-lait spots. ^26^ In our algorithm 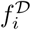 represents the numerator of the LR.

The denominator of the LR is the probability of the phenotypic feature if the proband does not have the disease (𝒟) in question. It would be difficult to calculate this for each of the 13,182 phenotypic abnormalities of the HPO in the general population, but we note that a tractable and realistic model for our purposes is that any proband being investigated by genomic diagnostics has some genetic disease. We can therefore calculate the denominator of the LR by means of the overall prevalence of HPO feature *h*_*i*_ in genetic diseases other than 𝒟. For instance, if 𝒟 and 13 of the 7622 other diseases in the HPO database are characterized by feature *h*_*i*_ and we assume an equal pretest probability for all diseases, then the probability of the proband having feature *h*_*i*_ if the proband is not affected by disease 𝒟 is the sum of the frequencies of *h*_*i*_ in the 13 diseases divided by 7622 (See the **Online Methods** for a detailed description of the algorithm).

Our algorithm takes as input a VCF file with genetic variants identified in an exome, genome, or gene panel experiment as well as a list of HPO terms that describe the phenotypic abnormalities observed in the proband. The algorithm returns a ranked list of candidate diagnoses each of which is assigned a posttest probability. Each of the HPO terms is conceived of as a diagnostic test, and a LR is calculated for each term representing the probability that a proband has the term in question if the proband has the candidate disease divided by the probability of the proband having the term if the proband does not have the candidate disease.

The current version of the HPO database comprises 7623 diseases, of which 5192 are associated with at least one gene (total disease-associated genes: 4025), and 2431 diseases are not associated with a gene. In contrast to previous approaches to phenotype-driven genomic diagnostics, ^1;2;27^ our approach includes diseases with no known associated disease gene in the differential. However, if a disease gene is known, then the genotype of the proband is also used as a diagnostic test in the LR framework. The LR is calculated for the observed genotype of the gene based on our expectation of observing one or two causative alleles according to the mode of inheritance of the disease and also the probability of observing called pathogenic variants in the gene in the general population. The individual LRs are multiplied to obtain a composite LR, which, together with the pretest probability of each disease, is used to calculate the posttest probability in order to rank the diseases.

### LIRICAL supports clinical interpretation with estimates of post-test probability and per-phenotype likelihood ratios

Fig. 1 illustrates our approach for a published proband with five characteristic features of Ataxia-pancytopenia syndrome (ATXPC; OMIM:159550): `Dysmetria`, `Babinski sign`, `Cerebellar atrophy`, `Dysarthria`, and `Ataxia`. ^28^ We additionally added the HPO term `High myopia` to simulate an unrelated (false-positive) finding that is not related to the underlying Mendelian disease. Exome sequencing was simulated in this example case by spiking a heterozygous variant in the causative gene for ATXPC, *SAMD9L*, into an otherwise “normal” VCF file. LIRICAL was then run on the combined phenotype and genotype data, and ranked ATXPC first out of the 7623 diseases in the HPO database. The graphical display of the results shown in Figure 1a indicates how much each feature contributed to the prediction. Figure 1b shows the second highest ranking candidate, Spinocerebellar ataxia, autosomal recessive 7 (SCAR7). SACR7 matches four of the five phentypic features that APS does. It scores lower because the match to the term `Dysmetria` was exact for ATXPC but in SPAR7 the closest match to `Dysmetria` was `Ataxia`, resulting in a lower LR (the HTML output of LIRICAL allows the user to browse the matching and approximate terms and their LRs by tool tips that appear when mousing over the bars that display the LR). The third candidate, Oculodental dysplasia (OMIM:164200) has two additional mismatching HPO terms, `Babinski sign` and `Cerebellar atrophy`, and is assigned a posttest probability of under 0.1%. LIRICAL thereby provides users both with an assessment of the degree to which any given phenotypic feature supports a diagnosis or argues against it, as well as an estimated posttest probability of the candidate diagnosis on the basis of the information provided. Users can remove terms deemed irrelevant (e.g., `High myopia`) and rerun the analysis. They can choose to concentrate detailed follow-up on candidate diagnoses with a high posttest probability.

**Figure 1:**
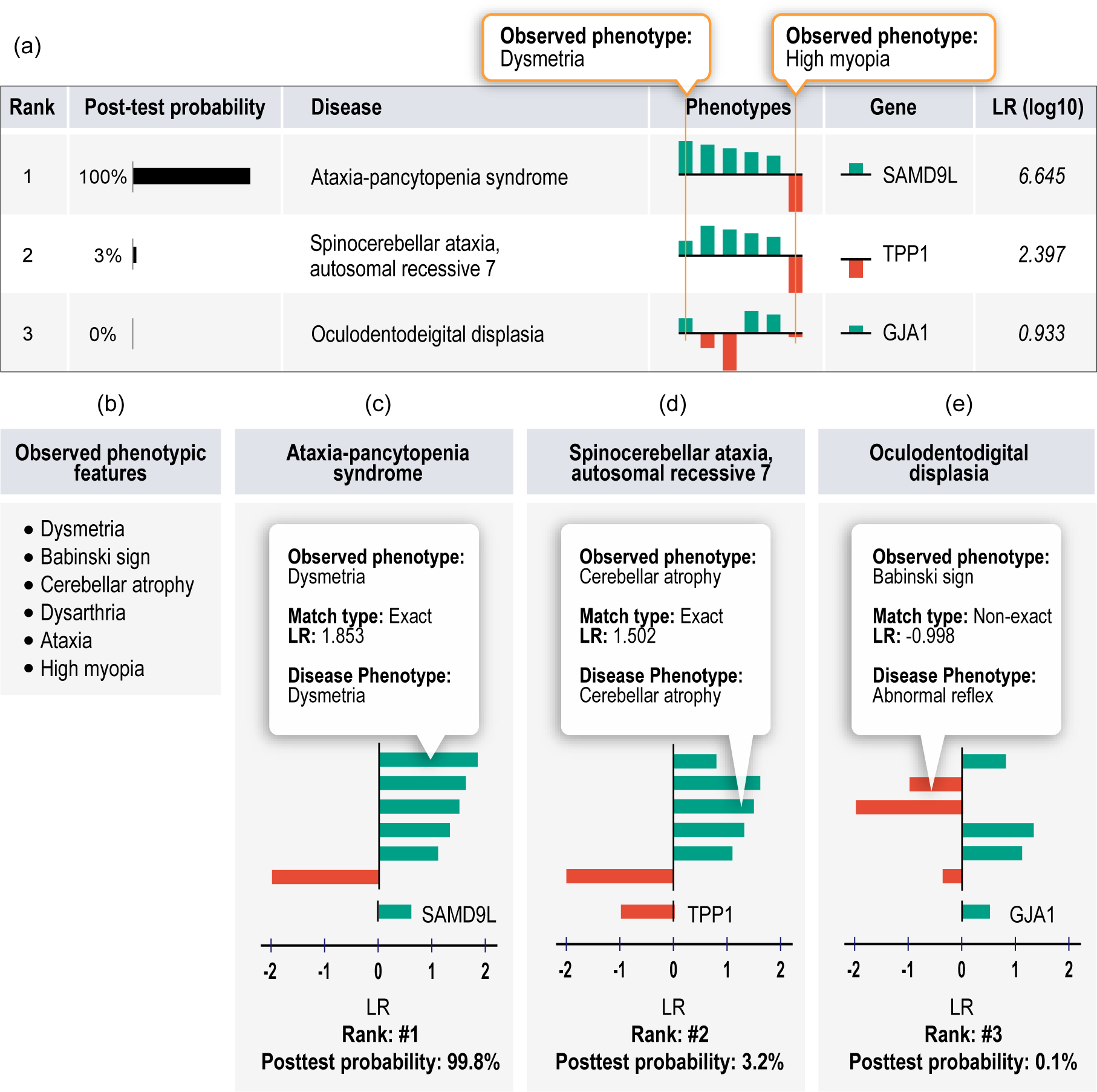
LIRICAL evaluation of simulated case of Ataxia-pancytopenia syndrome (ATXPC) For each candidate diagnosis with an above-threshold posttest probability, LIRICAL shows the contribution of each phenotypic feature and of the genotype to the final diagnosis. In this case, the data were extracted from a published case report on an individual with ATXPC, ^28^ and an additional unrelated term (`High myopia` was added to simulate the effect of noise. **(a)** LIRICAL provides a table of the top candidates with the posttest probability and a sparkline view of the contributions of each HPO term and the relevant genotype. **(b)** The observed HPO terms. **(c)** The correct diagnosis, ATXPC, is ranked in first place because of a good phenotype match and a positive LR for the heterozygous genotype for the causative gene *SAMD9L*. **(d)** The second candidate has many of the same phenotype matches, but the first query term `Dysmetria` matches exactly with Ataxia-pancytopenia syndrome and only approximately with the second candidate, spinocerebelllar ataxia, autosomal recessive 7. **(e)** The third candidate has a posttest probability close to zero because it has more mismatching or poorly matching query terms.

### LIRICAL achieves state of art performance and is robust to phenotypic and genotypic noise

We evaluated the performance of LIRICAL using several different approaches. Many previous studies simulated cases by choosing a certain number of HPO terms at random to simulate a patient (e.g., choosing 5 terms at random from the 56 terms that annotate Marfan syndrome in the HPO database). Phenotypic noise is simulated by adding a certain number of HPO terms at random from all available annotations (“noise terms”). In some cases, imprecision of clinical data entry is simulated by replacing the randomly chosen disease terms by parent terms. If studies simulate genomic analysis, then additionally a published disease-associated variant would be spiked into an otherwise normal VCF file. ^29–32^ However, this kind of simulation can be criticized because randomly chosen terms are unlikely to resemble terms that would be chosen in a real clinical encounter. In a real clinical encounter, the clinician may or may not be able to describe phenotypic abnormalities with the greatest possible detail. For instance, a general practicioner may diagnose reduced visual acuity, but the precise abnormality, say Y-shaped cataract, may only be observable by an ophthalmologist. Therefore, in real life situations, the different aspects of the phenotype of a patient may have been observed, recorded, or communicated at different levels of detail.

Our basic approach for this study was therefore to extract HPO terms and disease-causing variants from published case reports and to perform simulations with the original data as well as simulations in which varying types of phenotypic or genotypic noise were added. We tested the performance of LIRICAL using a collection of 384 case reports derived from the literature and curated using the GA4GH phenopacket format (Table 1). LIRICAL can be run with or without genetic data, and so we first compared it to Phenomizer, which exploits semantic similarity between query terms and diseases on the basis of clinical (but not genetic) data. ^29^ LIRICAL placed a total of 43.7% of cases in the top 3 ranks compared to 35.3% for Phenomizer (Supplemental Figure S2).

**Table 1:** Phenopackets used for evaluating the performance of LIRICAL. 384 phenopackets each describing a single published case report were derived from the literature by manual biocuration. See Supplemental Table S1 for details. Multiple modes of inheritance means that more than one mode has been described for the disease in question, e.g. inherited cataract associated with variants in *PITX3* can be inherited in an autosomal dominant or recessive fashion.

|  |  |
| --- | --- |
| **Cases** |  |
| total | 384 |
| **Diseases** |  |
| Median # cases per disease | 1 |
| Maximum # cases per disease | 19 |
| Autosomal recessive diseases | 203 |
| Autosomal dominant diseases | 128 |
| X chromosomal diseases | 10 |
| Multiple modes of inheritance | 43 |
| total | 262 |
| **Disease genes** |  |
| total | 259 |
| **HPO terms** |  |
| total over all cases | 1687 |
| Mean # HPO terms per case | 11.1 (median 9) |
| Mean # negated HPO terms per case | 2.71 (median 0) |

We then compared LIRICAL to Exomiser, which has shown state of art performance against other algorithms. ^31^ Exomiser currently ranks disease genes (combining all diseases associated with any given gene), and so for this comparison we recorded LIRICAL’s rank by gene. LIRICAL placed the correct gene in the first ranks in 80.7% of cases, compared to 77.3% for Exomiser. The percentages for placing the correct gene in the top 3 ranks were 92.9% for LIRICAL and 92.2% for Exomiser (Fig. 2b).

**Figure 2:**
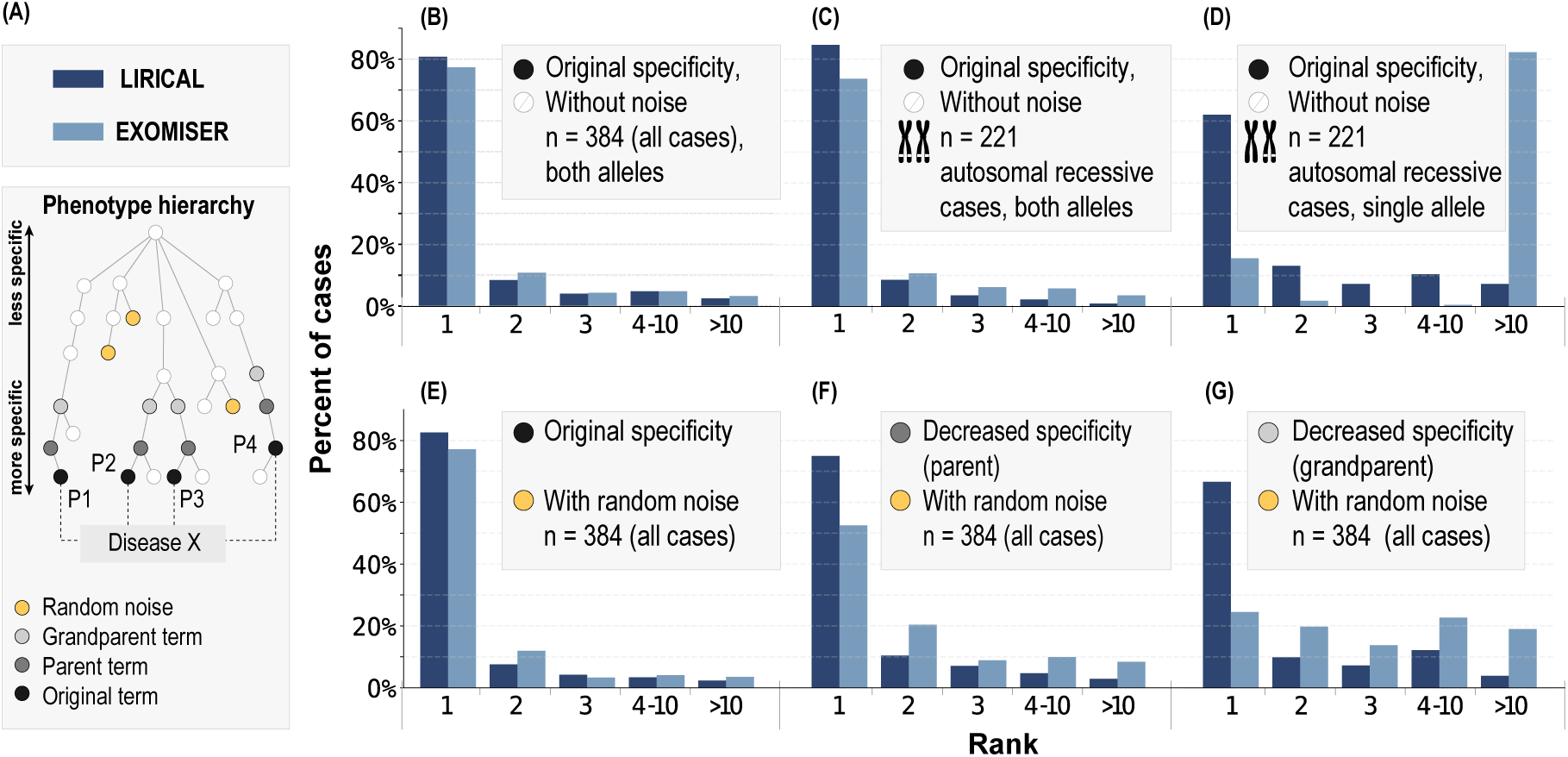
Evaluation of LIRICAL and Exomiser on 384 case studies. . The case studies were formated as Phenopackets (Table 1) and the diagnostic process was simulated by spiking disease-causing variants into a VCF file, which was passed together with phenotype data to LIRICAL and Exomiser. **(a)** Simulation approach. Random noise terms were added to some simulations, and in some cases, terms were replaced by their parent term or grandparent term to mimic imprecision in measuring or recording phenotypic abnormalities. **(b)–(g)** Results of simulations are shown with the X axis showing the rank assigned by LIRICAL or Exomiser to the correct disease gene, and the Y axis showing the percentage of cases in which the given rank was achieved. **(b)** Original data; **(c)** Performance on the subset of 221 autosomal recessive cases; **(d)** The same 221 autosomal recessive cases in which one of the two pathogenic alleles was removed. **(e)** Two random (“noise”) HPO terms are added to each case; **(f)** Original terms are replaced by a parent term and two noise terms are added; **(g)** Original terms are replaced by a grandparent term and two noise terms are added.

Diagnostic Next-Generation Sequencing (dNGS) data including exome, genome, and gene-panel investigations can be affected by many different kinds of noise. ^15^ The disease-causing variant may be missed, or in autosomal recessive conditions one of the two pathogenic alleles may fail to be detected. Phenotypic features unrelated to the Mendelian disease may be included in the analysis. On the other hand, phenotypic features associated with the disease may be observed or described imprecisely. LIRICAL was designed with a number of features that can help mitigate these kinds of noise.

We first compared the performance of both approaches in the presence of phenotypic noise Fig. 2a explains the obfuscations). Fig. 2e shows the performance if two random HPO terms are added to each case to simulate noise. Fig. 2f shows the effect of additionally replacing each of the original HPO terms with a parent term, and Fig. 2g shows the effect of additionally replacing each original term with a grandparent term. The latter two experiments simulate the effect of two different degrees of imprecision in the description of the clinical data (e.g., not entering a term such as `Zonular cataract` but instead entering its parent term `Cataract` or even grandparent term `Abnormality of the lens`). It can be seen that LIRICAL’s performance is better than Exomiser’s on this dataset and that LIRICAL’s performance degrades less in the presence of noise.

LIRICAL’s genotype LR does not apply a hard filter to candidates whose genotype does not match the expected genotype for some disease. In exome and genome sequencing, structural variants and point variants in GC rich exons may be missed, which can lead to only one of two pathogenic alleles being detected for an autosomal recessive disease. LIRICAL will rate such a genotype less highly than a pathogenic biallelic genotype, but will not filter out such candidates (Supplemental Figure S9). We therefore compared the performance of LIRICAL and Exomiser on the 221 autosomal recessive cases in our dataset. LIRICAL placed the correct candidate in first place in 84.6% of cases compared to 71.0% for Exomiser. If one of the two pathogenic alleles is removed, LIRICAL still placed the correct gene in first place in 62.0% of cases, compared to only 20.1% for Exomiser (Fig. 2c–d).

LIRICAL ranked 259 of 384 (67.4%) cases at a posttest probability above 0.5, and 287 cases (74.7%) were above a posttest probability of 0.05. The overall rankings as well as the posttest probability were robust to the addition of noise, deteriorating only slightly when two random terms were added per case, somewhat more if terms were replaced by more general parent or even more general grandparent terms, and falling to a mean of only 29.4% if all pathogenic alleles were omitted, and to 2.9% if all HPO terms were replaced by random terms (Fig. 3a). This suggests that LIRICAL assigns substantially mean lower posttest probabilities to candidates diseases for which by chance an apparently pathogenic variant is identified by dNGS but where there is no clinical match.

**Figure 3:**
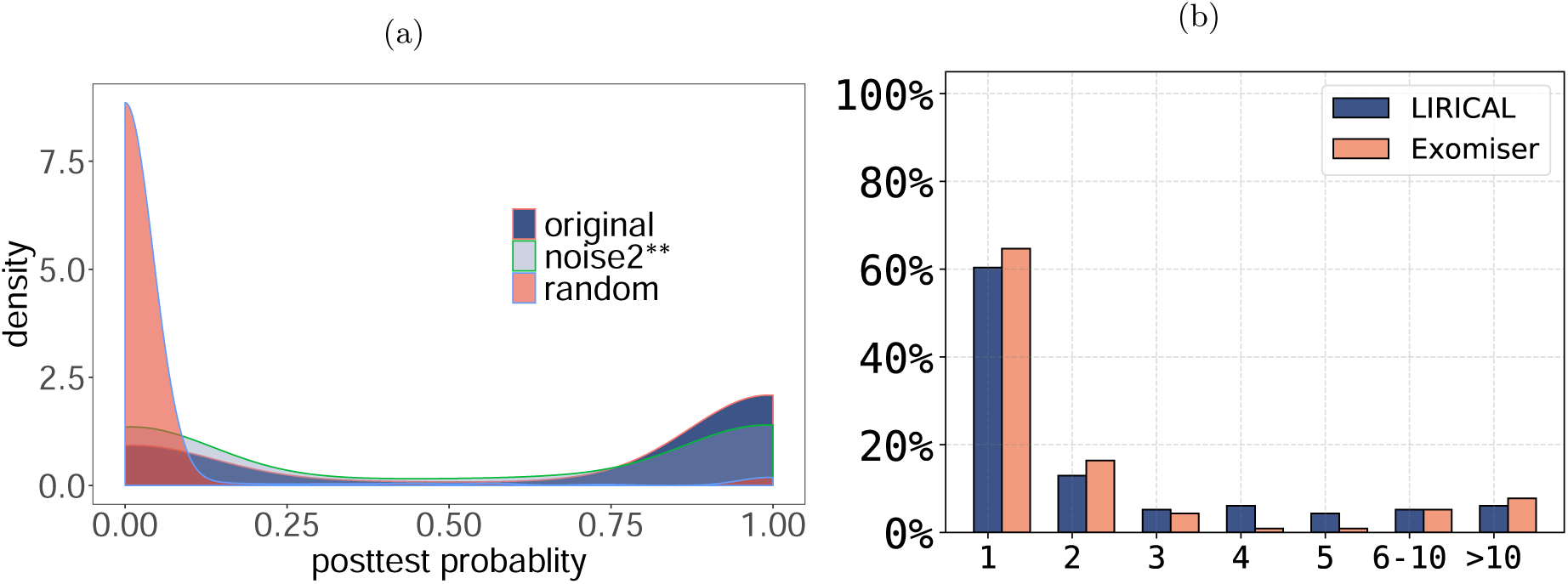
**(a)** The posttest probability of the correct diagnosis was calculated for each of the 384 phenopacket case reports (Original). Densities are shown for the original data (`original`; mean posttest probability, pp, 67.4%), `noise2**`: two random HPO terms were added and original terms were replaced by grandparent terms (mean pp: 50.3%), and `random`: All HPO terms were replaced by random terms (mean pp: 2.9%). Supplemental Figure S6 shows results for other perturbations. **(b)** Performance of LIRICAL and Exomiser on 116 solved singleton cases from the 100,000 Genomes project. The X axis shows the rank assigned by LIRICAL or Exomiser to the correct disease gene. The Y axis shows the percentage of cases in which the given rank was achieved.

Finally, we examined 116 solved singleton cases from the 100,000 Genomes Project. All cases were singletons with single-sample VCF files available. The diagnoses came from 89 different genes across a wide spectrum of rare disease areas (cardiovascular, ciliopathies, dermatological, dysmorphic and congenital abnormalities, endocrine, hearing and ear, metabolic, neurology and neurodevelopmental, ophthalmological, renal and urinary tract, rheumatological, skeletal, tumor syndromes). LIRICAL placed the correct gene in first place in 60.3% of cases, compared to 64.6% for Exomiser, and placed the correct gene in the top 5 ranks in 88.8% compared to 87.1% for Exomiser (Fig. 3b). This is an impressive outcome, considering that there may be a performance bias from Exomiser already being part of the 100,000 Genomes Projects diagnostic pipeline.

### Prioritization of genes associated with multiple diseases

Many Mendelian-disease related genes are associated with more than one disease (for instance, mutations in *FBN1* are associated with both Marfan syndrome and geleophysic dysplasia). In contrast to Exomiser, LIRICAL ranks diseases rather than genes (for an example, see Fig. 4). The by-disease ranking results for LIRICAL for the data in Fig. 2B are shown in Supplemental Fig. S3.

**Figure 4:**
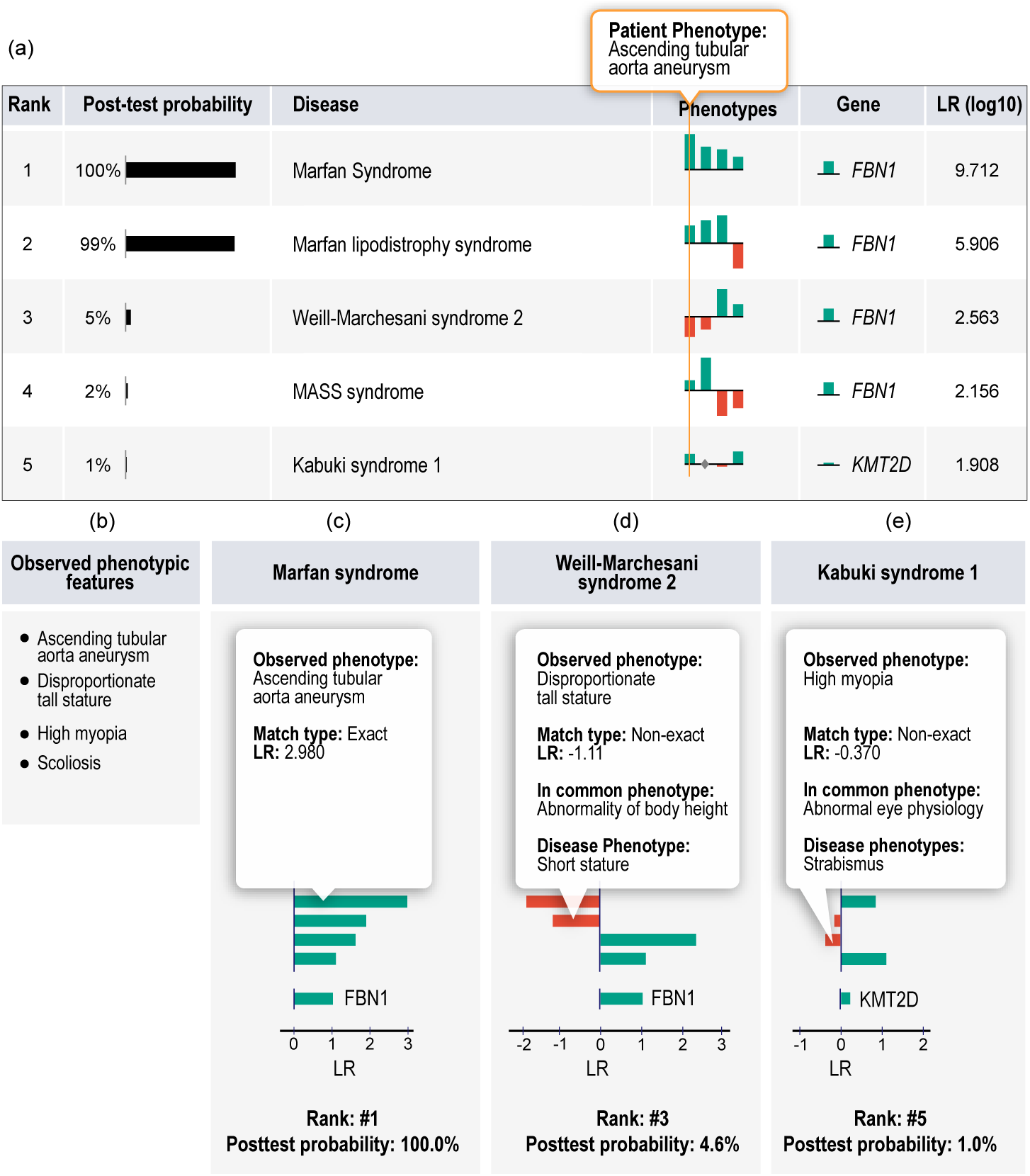
LIRICAL evaluation of simulated case with a pathogenic *FBN1* variant. Eight distinct diseases are associated with variants in the *FBN1* gene. LIRICAL prioritizes each disease separately, and in this case correctly placed Marfan syndrome at rank #1. Three other *FBN1*-associated diseases were placed in ranks 2–4. Clinical and molecular data were simulated according to patient 1 in Cao et al. ^33^. The HPO terms are shown in panel (b). The graphic shows LIRICAL’s summary table and three of the detailed LR plots for the candidates at ranks #1 (c), #3 (d), and #5 (e).

### Incorporation of ClinVar data and analysis of excluded phenotypic abnormalities

LIRICAL uses several heuristic algorithms to account for some challenges in the prioritization of genomic data. For instance, genes such as *TTN* have a high population frequency of variants predicted computationally to be pathogenic that are found in apparently healthy individuals. On the other hand, specific *TTN* variants are listed as pathogenic in ClinVar. ^34^ There is currently no approach that always correctly interprets pathogenicity of variants in such genes. In such cases, LIRICAL takes the approach of downweighting rare, predicted pathogenic variants without support in ClinVar, but heuristically assigns variants listed as pathogenic in ClinVar a LR score of 1000. In a simulated case of *TTN*-related dilated cardiomyopathy, LIRICAL correctly ranks a known pathogenic variant in first place but ranks a rare variant that is computationally predicted to be pathogenic but is listed in ClinVar as uncertain only in eigth place (Supplemental Fig. S12).

In clinical practice, the differential diagnostic process can occasionally be empowered by identifying phenotypic abnormalities that a patient does not have. In medical genetics, many diseases share a number of phenotypic features but differ with respect to one characteristic feature which presents in one disease but never presents in others. Such a feature can be very important for the differential diagnosis. For instance, Loeys-Dietz syndrome 4 is *not* characterized by `Ectopia lentis`, while the phenotypically similar disease Marfan syndrome is ^35^. LIRICAL uses a heuristic to downweight candidate diagnoses by a factor of 1000 if the candidate is explicitly annotated not to have a feature present in the query terms. 10 of the 380 phenopackets have excluded query terms (e.g., the individual does *not* have some HPO term) that support one candidate diagnosis (column 1 in Supplemental Table S3) but speak against another (column 2 in the table). In all cases, the correct diagnosis using the negated annotations was 1, with the mean posttest probability of 98.9%. If the negated query term was omitted, the average rank was 1.3, and the mean posttest probability was 72.6% (Supplemental Fig. S5). Supplemental Figure S10 shows an example of a differential diagnosis in which the omission of a negated term reduces the posttest probability of the correct diagnosis from 92.4% to 1.2% and changes the rank of the candidate from 1 to 2. To our knowledge, LIRICAL is the only HPO-based algorithm for genomic diagnostics that leverages information about excluded phenotypes in this way.

### Simultaneous analysis of molecularly elucidated and idiopathic diseases

Another feature of LIRICAL is a mode (`--global`) that ranks all candidates including diseases whose molecular etiology is unknown as well as diseases with a known associated gene in which no pathogenic variants were identified. This is a harder prediction problem because there are more candidate diseases, but it can prioritize diseases that would be missed by conventional approaches. For example, Arima syndrome is an autosomal recessive disease with no known disease-associated gene. LIRICAL prioritized it in first place in a simulated run, in which some clinically similar diseases such as Joubert syndrome failed to achieve a good score (Supplemental Fig. S11). LIRICAL placed the correct diagnosis in first place in 24.5% of cases compared to 1.0% for Exomiser, and placed the correct candidate in the top three ranks in 38.2% (1.0% for Exomiser). Overall, LIRICAL placed the correct candidate in the top ten ranks in roughly half of the cases (Supplemental Fig. S4).

## Discussion

Clinical decision support systems and genomic diagnostics have rapidly been gaining importance in recent years. The interpretability of computational predictions is of utmost importance in clinical settings for clinicians to efficiently and correctly integrate computational analyses into medical workflows, and even accurate black-box algorithms may not be appropriate in clinical settings. ^21;36;37^ The LIRICAL algorithm presented here adapts the LR framework that is widely used in the interpretation of clinical laboratory results. ^22;38;39^ To the best of our knowledge, the LR framework has not previously been used to support phenotype-driven genomic diagnostics. LIRICAL provides predictions of rare-disease diagnoses whose accuracy at par with that of previous state-of-art approaches such as Exomiser. ^27^ LIRICAL exhibits substantially better performance in the face of phenotypic and genotypic noise. Additionally, provides an estimated posttest probability of each candidate diagnosis and allows clinicians to evaluate the contribution of each individual phenotypic abnormality to each candidate diagnosis.

A LR indicates how many times more or less likely patients with the disease are to have that particular result than patients without the disease. A LR greater than 1 indicates that the result of the test is associated with the presence of the disease being investigated, while a LR less than one indicates the absence of the disease. The more the value of the LR deviates from 1, the stronger the evidence is for the presence or absence of disease. ^24^ In practice, the posttest probability can be used as an estimate of the quality of any diagnosis. The mean posttest probability estimated for the candidate at rank one for randomized data was close to zero, while the posttest probability of the correct diagnosis was about 67% for the case reports (Fig. 3). In some cases, however, the correct candidate was placed at rank one but received a low posttest probability. Future improvements in the quality and comprehensiveness of HPO annotations as well as in the computational assessment of variants may lead to improved ability of LIRICAL to estimate posttest probabilities.

LIRICAL can analyze an exome in less than a minute on a typical laptop computer. We identified 14 other tools for phenotype driven analysis of diagnostic exome or genome data. None of these tools was both up to date and available for executation on the command line, which would have enabled testing of the total of 1978 original or obfuscated cases from the phenopackets and the 116 GEL cases (Supplemental Table S2).

In addition to having a performance that is comparable to that of other state of art tools such as Exomiser, LIRICAL provides users with interpretable results that can be used to guide clinical actions. For instance, large-scale disease sequencing projects such as the 100,000 Genomes Project often have hundreds or thousands of unsolved cases. LIRICAL can be run on collections of unsolved cases, and the posttest probability of the highest ranked candidates could be used as a critierion to decide whether to subject a case to detailed reanalysis.

LIRICAL’s assessment of the contribution of individual phenotypic abnormalities can also be useful in many ways. For instance, in practice, patients with genetic diseases may present with a mix of signs and symptoms that are related both to an underlying Mendelian disorder and also may have unrelated (coincidental) findings. If a core set of phenotypes and a genotype strongly support a candidate diagnosis but some features do not, clinicians may consider whether alternate explanations for the non-contributory features are plausible according to their clinical judgment. For instance, features such as Myopia, Scoliosis, and Gastroesophageal reflux are relatively common in the general population and may therefore occur in persons with genetic disease as coincidental findings. Clinical judgment would be necessary to evaluate each term. For instance, myopia (short-sightedness) is relatively common in young adults, but the presence of high myopia in a toddler is more likely to be a clinical finding that is important for the differential diagnostic workup.

LIRICAL takes as input a list of HPO terms and can be run with or without an associated VCF file with genetic variants. The Java implementation of LIRICAL presented here assumes an equal pretest probability for each of the diseases under consideration (e.g.,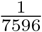 for the 7596 diseases currently represented in the HPO database). This is a reasonable approach to the analysis of Exomes in a setting such as the 100,000 Genomes Project where we speculate that rarer genetic diseases are more likely to be analyzed than common, more easily recognized genetic diseases. However, in other settings LIRICAL could be used with other values for the pretest probability. For instance in general care settings, the rare-disease prevalence data from Orphanet could be used. ^40^

LIRICAL is implemented as a standalone Java desktop application that can be installed in less than an hour. LIRICAL is freely available for academic use and source code can be downloaded from https://github.com/TheJacksonLaboratory/LIRICAL.

## Methods

### Data sources

The `hp/releases/2019-09-06` version of the Human Phenotype Ontology (`hp.obo`) was used for the analysis described here (http://purl.obolibrary.org/obo/hp.obo). HPO annotations (HPOA) were downloaded on October 16, 2019 from http://compbio.charite.de/jenkins/job/hpo.annotations.current/lastSuccessfulBuild/artifact/misc_2018/phenotype.hpoa.

### Likelihood ratio

The likelihood ratio (LR) is defined as the probability of a given test result (x) in a patient with a target disorder 𝒟 divided by the probability of that same result in a person without the target disorder (¬𝒟):

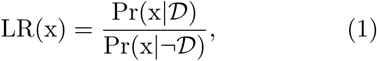

Pr(x|𝒟) is the sensitivity (true positive rate) of the test, i.e., the proportion of individuals with disease 𝒟 who are correctly identified. The specificity or true negative rate is the proportion of individuals without disease 𝒟 who are correctly identified as unaffected, i.e., Pr(¬x|¬𝒟). Therefore, the LR can be expressed as

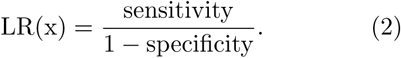

The definition of the LR can be extended to multiple tests. ^22^ Suppose *X* = (*x*_1_, *x*_2_, …, *x*_*n*_) is an array of *n* test results. Under the assumption that the tests are independent, LR(X) is defined as:

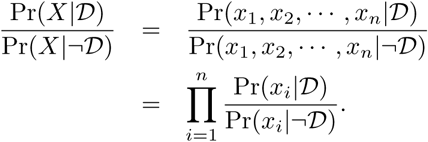

The *posttest probability* refers to the probability that the patient has a disease given the information from test results *X* and the pretest probability of the disease. The posttest probability can be calculated as

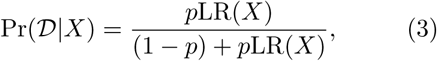

where *p* is the pretest probability of 𝒟. Depending on the cohort, the pretest probability can be defined as the population prevalence of the disease or by some other estimate of the frequency of the disease in the cohort being tested.

### Likelihood ratio for phenotypes

The signs and symptoms and other phenotypic abnormalities of probands being investigated by this approach are represented using terms of the Human Phenotype Ontology (HPO), which provides a structured, comprehensive and well-defined set of 14,813 classes (i.e., terms; September 2019 release) describing human phenotypic abnormalities ^13;41–43^. We model the clinical encounter that results in a set of *n* phenotypic observations encoded as HPO terms *h*_1_, *h*_2_, …, *h*_*n*_. The LR of each phenotype term with respect to a specific disease 𝒟 is defined as:

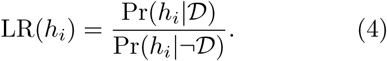

We assume that the tests are independent and the LR of the *n* HP terms are obtained from the product of the individual ratios.

### The probability of having phenotypic abnormality *h*_*i*_ given a disease 𝒟

We first explain how we define the numerator of equation (4) based on the relationship of term *h*_*i*_ to the set of phenotype terms to which disease 𝒟 is annotated (Supplemental Figure S1). We distinguish four cases.

#### (i) *h*_*i*_ is identical to one of the terms to which 𝒟 is annotated

In this case, we define 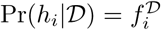, that is, the frequency of the phenotypic feature *h*_*i*_ amongst individuals with disease 𝒟. For instance, if the disease model for 𝒟 is based on a study in which 7 of 10 persons with 𝒟 had *h*_*i*_, then 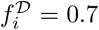. If no information is available about the frequency of *h*_*i*_, then by default we define 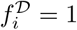.

#### *(ii) h*_*i*_ is an ancestor of one or more of the terms to which 𝒟 is annotated

Because of the annotation propagation rule of subclass hierarchies in ontologies ^44^, 𝒟 is implicitly annotated to all of the ancestors of the set of annotating terms. For instance, if the computational disease model of some disease 𝒟 includes the HPO term `Polar cataract` (HP:0010696) then the disease is implicitly annotated to the parent term `Cataract` (HP:0000518) (to see this, consider that any person with a polar cataract can also be said to have a cataract). By extension, this is also true of more distant descendants of the term. We therefore define the probability of a term *h*_*i*_ (e.g., `Cataract`) that is an ancestor of any term *h*_*j*_ (e.g., `Polar cataract`) that explicitly annotates disease 𝒟 as

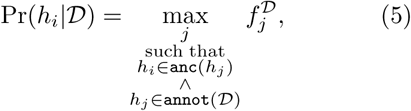

where `anc`(*h*_*j*_) is a function that returns the set of all ancestors of term *h*_*j*_ and `annot`(𝒟) is a function that returns the set of all HPO terms that explicitly annotate disease 𝒟. In words, the probability of *h*_*i*_ in disease 𝒟 is equal to the maximum frequency of any of the descendants of *h*_*i*_ that directly annotate disease 𝒟.

#### (iii) *h*_*i*_ is a child term of one or more of the terms to which 𝒟 is annotated

In this case, *h*_*i*_ is a descendant (i.e., specific subclass of) some term *h*_*j*_ of 𝒟. For instance, disease 𝒟 might be annotated to `Syncope` (HP:0001279), and the query term *h*_*i*_ is `Orthostatic syncope` (HP:0012670), which is a child term of `Syncope`. In addition, `Syncope` has two other child terms, `Carotid sinus syncope` (HP:0012669) and `Vasovagal syncope` (HP:0012668). According to our model, we will weight the frequency of `Syncope` in disease 𝒟 (say, 0.72) by 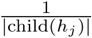, where child(*h*_*j*_) is the set of child terms of *h*_*j*_ (so in our example, we would use the frequency 0.72 *×* 1/3 = 0.24). In our implementation, only the direct children of a disease-associated term *h*_*j*_ are considered. The maximum LR is taken across all disease-associated terms.

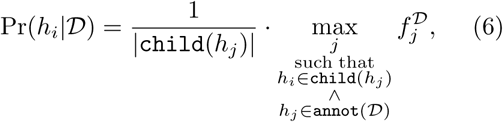

where `child`(*h*_*j*_) refers to the set of direct descendants (child terms) of HPO term *h*_*j*_.

#### (iv) *h*_*i*_ and some term to which is 𝒟 annotated have a non-root common ancestor

In this case, *h*_*i*_ is not a child term of any disease term *h*_*j*_ and no disease term *h*_*j*_ is a descendant of *h*_*i*_.

If this is the case, then we find the closest common-ancestor (denoted *h*_*ca*_ in the following). Noting that *h*_*ca*_ may have a zero or very small frequency in disease 𝒟, we define the LR using the following heuristic:

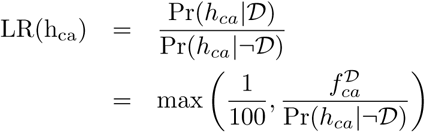

Because the common ancestor is higher up in the HPO hierarchy, the LR tends to be lower and sometimes substantially lower for features with a high frequency across the HPO corpus (with a corresponding low value for Pr(*h*_*ca*_ | ¬ 𝒟). Therefore, in order to avoid a single term having an excessive influence on the final result, the LR is taken to be at least 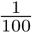,

#### (v) *h*_*i*_ does not have any non-root common ancestor with any term to which 𝒟 is annotated

In this case, *h*_*i*_ does not affect the same organ system as any of the annotations of 𝒟. A heuristic small value of 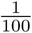 is assigned.

### The probability of having a phenotypic abnormality *h*_*i*_ that is explicitly excluded from disease 𝒟

In the HPO annotation resource, each disease is represented by a list of HPO terms that characterize it together with metadata including provenance, and in some cases, frequency and onset information ^13^. Some diseases additionally have explicitly excluded terms (there are a total of 921 such annotations in the September 2019 release of the HPOA data). These annotations are used for phenotypic abnormalities that are important for the differential diagnosis. For instance, Marfan syndrome and Loeys-Dietz syndrome share many phenotypic abnormalities. The feature `Ectopia` `lentis` (HP:0001083) is characteristic of Marfan syndrome but is not found in Loeys-Dietz syndrome ^45^. The LR for such query terms is assigned an arbitrary value of 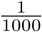, i.e., the ratio for a candidate diagnosis is reduced by a factor of one thousand if an HPO term is present in the proband that is explicitly excluded from the disease.

On the other hand, if the query includes a negated term that is explicitly excluded in the disease, then the opposite value is assigned, i.e., the ratio for a candidate diagnosis is increased by a factor of one thousand if an HPO term is present in the proband that is explicitly excluded from the disease.

### The probability of having phenotypic abnormality *h*_*i*_ if disease 𝒟 is not present

The denominator of equation (4) specifies the probability of the test result given that the proband does *not* have some disease 𝒟. This would be difficult to calculate for the general population for the same reasons as those described above. However, we can estimate this probability if we assume that all persons being tested have some (unknown) Mendelian disorder by simply summing over the overall frequency of a feature in the entire HPO corpus (with *N* diseases).

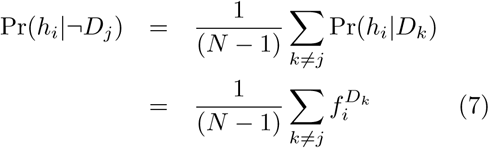

Equation (7) would need to be calculated separately for each of the *N* diseases, but noting that we are summing over a relatively large number of diseases (7623 in September, 2019) in the complete HPO database of rare diseases, we use the following approximation that allows us to precalculate Pr(*h*_*i*_|¬*D*_*j*_) for an arbitrary disease *D*_*j*_.

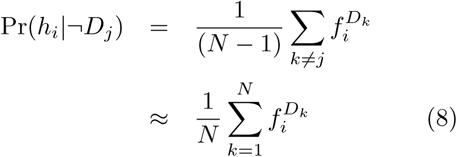

### Likelihood ratio for genotypes

Our model of predicting the relevance of any given genotype makes use of the following concepts. There is a true but unobservable pathogenicity, defined as a deleterious effect of a genetic variant on the biochemical function of a gene and the gene product it encodes that leads to disease. We can estimate the pathogenicity of a variant on the basis of a computational pathogenicity score that ranges from 0 (predicted benign) to 1 (maximum pathogenicity prediction). Our model posits two distributions that allow us to calculate the likelihoods of an observed genotype given that the sequenced individual has the disease (𝒟) as compared to the situation in which the individual does not have the disease in question and the variants originate from population background (ℬ). We will use the pathogenicity score of the Exomiser, which calculates a score for any variant in the coding exome or at the highly conserved dinucleotide sequences at either end of introns ^27^. We will use the estimated population frequencies of variants from the gnomAD ^46^ resource, which is incorporated into the Exomiser database to calculate the background distribution (version 12.1.0 was used for the analysis reported here).

Our model depends on the assumed mode of inheritance of the disease; we will begin our explanation with autosomal dominant (AD) diseases. We are interested in the ratio of an observed genotype (𝒢) given that it is disease-causing (i.e., the sequenced individual has disease 𝒟) or not (i.e., the sequenced individual does not have disease 𝒟). Assume we observe *n* variants (*v*_1_, *v*_2_, …, *v*_*n*_) in gene *g*, and have calculated their pathogenicity score as *s*(*v*_*i*_) for *i* ∈ {1, …, *n*}. For simplicity, we will assume that the variants have been arranged such that *s*(*v*_1_) ≥ *s*(*v*_2_) ≥ … ≥ *s*(*v*_*n*_).

We first note that 98.9% of the pathogenicity scores of variants classified as pathogenic in ClinVar ^34^ are assigned a pathogenicity score of 0.8 or more by Exomiser (**Supplemental Figure S7**). For the purposes of assessing and scoring candidate variants, we therefore divide the score distribution into two bins 𝒩 and 𝒫, with bin 𝒩 representing the *predicted non-pathogenic* bin and having a range of pathogenicity scores of [0, 0.8), and bin 𝒫 representing the predicted pathogenic bin with pathogenicity scores of [0.8, 1]. Although in reality there is no strict division in pathogenicity scores between neutral and disease-causing variants, we will use the binning as a way of down-weighting variants in genes that often show predicted pathogenic variants and tend to be frequently found as false positives in exome sequencing results, such as many mucin and HLA genes ^47^.

We model the expected counts of observed alleles in bin as Poisson distributions, using separate distributions for the case that a variation in a given gene is disease-causing or not. In this context, a Poisson distribution gives the probability of observing *k* variants in a gene, based on a rate parameter *λ* that represents the expected number of variants.

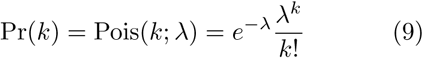

For an autosomal dominant disease associated with pathogenic variants in gene *g*, we expect one heterozygous disease causing variant, and so 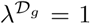; for autosomal recessive diseases, 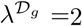. We can estimate the probability of observing a variant 𝒫 in bin in a gene *g* that is not related to the disease based on the frequency of such variants in the general population; we denote this probability as 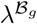. Different genes have different distributions of predicted pathogenic variants in the general population. If a gene has a low frequency of predicted-pathogenic variants in the general population, then the observation of a predicted-pathogenic variant in a diagnostic context may be more likely to be a true-positive disease-causing variant ^48^. We calculate 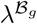 for each gene *g* based on available population frequency data from the gnomAD ^46^ resource by summing up the frequencies of individual variants under the independence assumption.

In detail, the frequency (if available) of each variant is taken from each of the following populations: African/African American (`GNOMAD_E_AFR`), Admixed American (`GNOMAD_E_AMR`), Ashkenazi Jewish (`GNOMAD_E_ASJ`), East Asian (`GNOMAD_E_EAS`), Finnish (`GNOMAD_E_FIN`), Non-Finnish European (`GNOMAD_E_NFE`), and South Asian (`GNOMAD_E_SAS`). For the analysis reported here, the average frequency in all populations is calculated. We note that this approach may overestimate the overall frequency of variants per exome/genome, but nonetheless we can use it as a heuristic to downweight genes commonly found to have predicted pathogenic variants in the population (e.g., Supplemental Table S5), as we will show below.

We denote the function that returns the predicted pathogenicity of a variant as `path` and the function that returns the average population frequency of a variant as `freq`. This parameter is calculated separately for each gene. We represent the fact that variant *i* is assigned to gene *g* as *v*_*i*_ ∈ *g*.

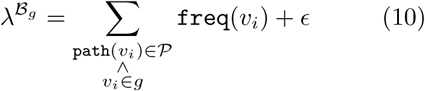

The parameter 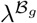 is thus the expected count of variants in gene *g* whose pathogenicity score is in bin 𝒫. A small number (*ϵ* = 10^−5^) is added to the sum to avoid division by zero in subsequent steps because some genes did not display any variants in bin 𝒫 in the population data.

For a gene associated with an autosomal dominant disease, the calculation proceeds as follows. Assume we are evaluating disease 𝒟 which is associated with mutations in gene *g* and that there is one predicted-pathogenic variant *v*^*t*^ in bin 𝒫 and there are *k* other predicted non-pathogenic variants in bin 𝒩. The model assumes that any variants in bin 𝒩 are unrelated to the disease and have the same probability whether or not gene *g* is causally related to the disease. The genotype observed for gene *g* is symbolized as `gt`(*g*).

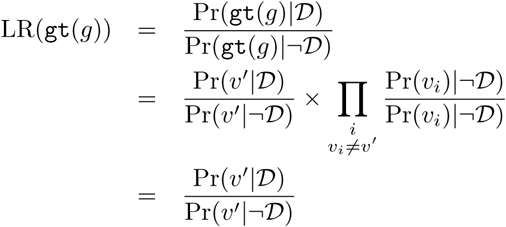

We model the process by which a variant or variants lead to disease by a compound distribution. A Poisson distribution models the number of variants observed whose pathogenicity score is in bin 𝒫, and a Bernoulli distribution with parameter *p* = *s*(*v*^*t*^) determines the probability that the allele is disease causing. Thus, let {**X**_*n*_} be a sequence of mutually independent random variables each of which can take on the value of 0 (for *not disease-causing*) or 1 (for *disease-causing*). The sum of *N* such variables is *S*_*N*_ = *X*_1_ + *X*_2_ + … + *X*_*n*_, and thus *S*_*N*_ represents the count of truly pathogenic alleles (we expect *S*_*N*_ = 1 for autosomal dominant and *S*_*N*_ = 2 for autosomal recessive diseases).

This leads to the compound distribution:

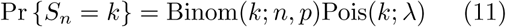

It can be shown that this is equivalent to a Poisson distribution with parameter *λp* ^49^. Therefore, to calculate the LR, we substitute the parameters 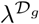 and 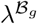 as well as *p* = *s*(*v*_*i*_).

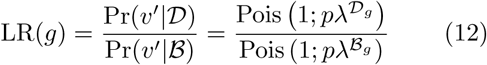

This will have the effect of favoring genes with a single variant in bin 𝒫 that has a maximal pathogenicity score (*p* = *s*(*v*^*t*^) = 1) and that has a minimal frequency of bin 𝒫 variants in the population. If this is the case, then 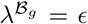 and we can calculate the LR using equation (9):

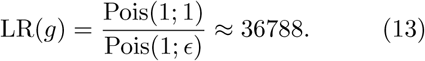

The procedure for autosomal recessive diseases is analogous, except that 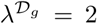. In the case that gene *g* is causative for the disease in the individual being sequenced, then we expect to find two alleles (which will be identical in case of a pathogenic homozygous variant and distinct in the compound heterozygous case). The two alleles in bin 𝒫 with the highest pathogenicity score are chosen for analysis. Let *s*_*avg*_ denote the mean of the pathogenicity scores of the two variants observed in gene *g* that have the two highest pathogenicity scores, i.e., *s*_*avg*_ = 0.5 *·* (*s*(*v*_1_) + *s*(*v*_2_)). Then,

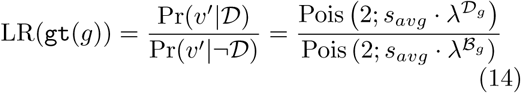

This will have the effect of favoring genes with two pathogenic alleles (homozygous or compound heterozygous) in bin 𝒫, which have a maximal pathogenicity score (*s*(*v*^*t*^) = 1) and that has a minimal frequency of bin 𝒫 variants in the population (in this case, 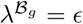 and LR(*g*) ≈ 3, 678, 831, 200, but this value is not seen in practice).

Noting that in males, hemizygous variants on the X chromosome are called as homozygous by current variant-calling software, we set 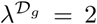 for both recessive and dominant X-chromosomal diseases.

### Genotype likelihood ratio: Special cases

#### (i) No variants at all found in gene *g*

If the molecular basis of a disease is known to be mutations in a gene *g*, but no bin variants or no variants at all are found in that gene, then a LR of 1/20 is assigned for autosomal dominant diseases, reflecting an estimation that the probability of missing a pathogenic variant if one is present is about 5%. For autosomal recessive diseases, we will estimate the probability at 0.05 *×* 0.05 = 0.0025.

The intuition for this step is that some down-weighting should be performed if no candidate variant is found in a gene but given the presumed high prevalence of false-negative results in exome/genome sequencing, it would not be desirable to radically downweight otherwise strong candidates.

#### (ii) Clinvar-pathogenic variant(s) found in gene *g*

ClinVar ^34^ makes use of the American College of Medical Genetics and Genomics and the Association for Molecular Pathology standards for the interpretation of a variant as `pathogenic` (i.e., causative of a disease) ^50^. Denote the count of ClinVar `pathogenic` alleles as *c*. If *c* = 2 for an autosomal recessive diseases, then a heuristic LR of 1000^2^ is assigned. If *c* = 1 for an autosomal dominant disease, then a heuristic LR of 1000 is assigned. If the *c* does not match the count of pathogenic alleles that would be expected for the mode of inheritance, then a heuristic LR of 1000 is assigned.

This heuristic means that if a ClinVar `pathogenic` variant is found even in a gene such as *TTN* that is characterized by a high frequency of predicted pathogenic variants in the population, then this is taken as being supportive of a diagnosis associated with variants in the gene.

#### (ii) Heuristic for genes with many variants

Some genes commonly harbor variants in the general population that are predicted as pathogenic by bioinformatic software. LIRICAL uses the background score to assess this. The background score ranged from 0 to 20.7 (for *MUC4*). Numerous disease-associated genes displayed scores over 1.0, including for example *TTN* with a score of 9.5. According to our model, it is not surprising to observe a predicted pathogenic variant in a gene such as *TTN*, whether or not the gene is associated with the disease being investigated in any particular case. LIRICAL downweights the LR for genotypes in these genes if predicted pathogenic variants are found in a VCF file, because such variants are commonly encountered as false positive findings. ^15^ It does so by limiting the value of 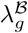 to be at most the observed count of predicting pathogenic variants, *c*_*path*_, in cases where 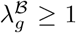 (If the observed called-pathogenic variant count is much higher, the probability calculated by the Poisson distribution will be very low).

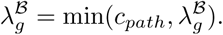

For instance, if one predicted pathogenic variant is identified in *TTN*, this scheme would lead to a LR of one – the observation of the predicted pathogenic variant in this gene neither adds to nor detracts from the probability of the differential diagnosis (we treat known disease-associated variants in ClinVar differently, see above).

### --global setting for genotype likelihood ratio

Our approach has two options for dealing with genes in which no predicted pathogenic variants are obeserved. With the default option, LIRICAL will remove the genes and the diseases they are associated from further analysis. This may be most appropriate if the goal of analysis is to demonstrate the genetic etiology of a disease.

If the `--global` option is chosen, LIRICAL ranks all diseases (including those with and without known associated disease genes) according to the posttest probability. In this case, if a disease has no associated disease gene, the LR is calculated from the phenotype evidence alone. Our procedure is designed to work whether or not genetic evidence is available to support a candidate diagnosis. If for instance, the individual being sequenced is affected by a Mendelian disease for which the causative genes have not yet been identified, then if there is a good phenotypic match, ideally the analysis procedure would include the disease in the overall results. Therefore, we omit the genotype score from the overall LR for Mendelian diseases in the HPO database that have a currently unclarified molecular basis.

### Combined genotype-phenotype likelihood ratio score

Our procedure takes as input a VCF file and a list of HPO terms representing the set of phenotypic abnormalities observed in the individual being sequenced. For each of the *∼*4,000 Mendelian diseases in the HPO database for which the causative disease gene has been identified, all bin `A` variants are extracted and their average pathogenicity score is calculated. The LRs are calculated for each phenotypic feature as described above. The final LR for some disease 𝒟 is then

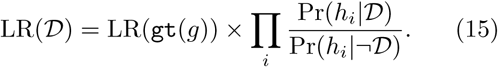

### Ranking candidates

Our approach calculates the LR of equation (15) for each disease represented in the HPO disease database (*n* = 7623 in the 9/2019 release). The diseases are then ranked according to the posttest probability.

### Visualization

The results of analysis are displayed here by showing bars whose magnitude is proportional to the decadic logarithm of the LRs of each tested feature. Features that support the differential diagnosis are shown in green and directed to the right of a vertical line in the center of the plot, and features that speak against the differential diagnosis are shown in red and directed to the left.

### Evaluation

We curated HPO terms from 384 published case reports (Table 1 and Supplemental Table S1). We chose case reports in which the causative mutation had been identified so that we could perform simulations with and without a simulated exome. We compared the results of simulation with the original data and also performed various types of obfuscation to assess the influence of noise on the performance of LIRICAL and Exomiser, adding varying degrees of phenotypic or genotypic noise (Supplemental Table S4).

A comparison of LIRICAL and Exomiser was also performed for 116 solved cases from the 100,000 Genomes Project where detailed clinical phenotype data in the form of HPO terms had been collected. All cases were singletons with single-sample VCF files available. The diagnoses came from 89 different genes across a wide spectrum of rare disease areas (cardiovascular, ciliopathies, dermatological, dysmorphic and congenital abnormalities, endocrine, hearing and ear, metabolic, neurology and neurodevelopmental, ophthalmological, renal and urinary tract, rheumatological, skeletal, tumour syndromes).

### Implementation

LIRICAL is implemented as a Java application. It is written in Java 1.8 and compiles under Java 11. An executable and source code can be downloaded from the GitHub page at https://github.com/TheJacksonLaboratory/LIRICAL and detailed documentation is available at the readthe docs page (https://lirical.readthedocs.io/en/latest/). LIRICAL is freely available for academic use.

## Data Availability

No new data was generated for this manuscript. Source code is available at https://github.com/TheJacksonLaboratory/LIRICAL

## FUNDING

This work was supported by internal funding of the Jackson Laboratory. Additional support was provided by the National Institutes of Health (NIH) Office of the Director (OD); The Monarch Initiative [1R24OD011883]. The UNC Biocuration core was supported by NHGRI U41HG009650.

## Notes

### Competing Interest Statement

Commercial users will be required to pay a license fee.

